# A Study On Clinical Profile And Semiology In Complex Partial Seizures And Its Radiological Correlation

**DOI:** 10.1101/2021.05.26.21257846

**Authors:** B. Nandini Priyanka, S. Elongovan, M. Thangaraj

**Affiliations:** Department of Neurology, Thanjavur Medical College - Thanjavur. 613004

**Author notes:** **Corresponding author:** Dr. S. Elongovan, Associate professor of neurology, Department of Neurology, Thanjavur Medical College-Thanjavur. 613004.

**Keywords:** Complex partial seizures, atypical febrile seizures, automatisms, aura, medial temporal lobe sclerosis

## Abstract

**Background:** Epilepsy is the second most common and frequently encountered neurological disorder which poses huge threat to known healthcare systems worldwide also causing financial, socio economic burden to the community. Complex Partial Seizures are a form of focal epileptic seizures that may impair consciousness.

**Aim and objectives:** Our motivation for the study was to understand the extent of patients with complex partial seizures associated with medial temporal lobe sclerosis.

**Materials and methods:** We performed a cross sectional study about patients with complex partial seizures in Thanjavur medical college and hospital about their clinical profile and neuro-radiological correlation.

**Statistical analysis and results:** Through our multimodal study with EEG, MRI on N=118 (female / male, age range, Most common age group-10-20 years, mean age of 23 years, SD-14 years, 66% males), we observed that atypical febrile seizures and fever provoked seizures has more association (18%) to complex partial seizures and to medial temporal lobe sclerosis in comparison to 12% in an earlier study.

**Conclusion:** We believe this study summarizes the complex partial seizure features, origin, and their link to Medial Temporal lobe Sclerosis in our subject pool from Thajavur, India.

**Limitation:** There are some limitations to our study, especially with no video EEG monitoring and no invasive EEG recording. We aim to improve them in our future studies.

## Introduction

Epilepsy is the second most common and frequently encountered neurological disorder which poses huge threat to known healthcare systems worldwide. Although it had been existing for more than 2000 years (as described by Hippocrates^1^) a recent study shows 70 million people were affected worldwide and particularly in India more than 12 million are with Epilepsy –People With Epilepsy (Estimated median prevalence 1.54% for Rural and 1.03% for Urban population and so, it draws the serious attention of medical community and necessary reviews are being undertaken. The incidence rate reported from India (0.2-0.6%) per 1000 population are comparable with developed countries and lower than most of the developing countries (1.0-1.9%)per 1000^2^. Thus it is evident that there exists a wide Rural—Urban gap for Incidence of Epilepsy with likely hotspots in Urban area due to migrant Rural population.

Complex partial seizures, preferably called as “focal impaired awareness seizure” or “focal onset impaired awareness seizure”, refer to focal seizures that start in one hemisphere of the brain and are associated with impairment in consciousness. International League Against Epilepsy (ILAE) 2017 classification has categorized seizures^3^ based on 1) the location of seizure onset, 2) level of awareness during a seizure and features such as automatisms, which is a coordinated, repetitive motor activity, often resembling a voluntary movement by undertaken without volition—it gets classified as focal onset seizures with types such as orofacial, unilateral or bilateral limb based manual, pedal movements, preservative, vocal, verbal, sexual, etc., and subjects may either have awareness or possess impaired awareness. Other features include aura, which is a subjective sensation that concurs with seizures and presents as a predictor. We present a study of the clinical profile of complex partial seizures, the nature of automatisms, and the lateralizing and localizing values of automatisms along with semiology features, its radiological correlations.

Fever involves elevation of temperature that influences many neurons in the brain to augment neuronal firing, that in turn can facilitate seizures. Some studies suggest that the underlying cause for epileptogenesis can promote inflammatory responses as well and the seizures could be an interplay between inflammation and fever. Some other studies doesn’t favour the argument.

There are some debates on the link between complex seizures and sclerosis (Mesial Temporal Sclerosis, MTS). Many studies generally link atypical febrile seizures to temporal sclerosis while some doesn’t^4–8^. We also aim to understand this link in our cohort of subjects through this study.

## Methods

We recruited N=118 (female / male, age range, Most common age group-10-20 years, mean age of 23 years, SD-14 years, 66% males) over the duration of 6 months. The subjects consented to the observational study, followed protocol as directed by TamilNadu Dr MGR medical university, Tamil Nadu, India.

Inclusion criteria: Patients with automatisms that progress to motor or non-motor features, with aura, impaired awareness, as predictive markers. Exclusion criteria include presentation of clinical features that are not suggestive of complex partial seizures, or seizures with multiple semiology other than complex partial seizures.

We performed EEG, CT, MRI on these subjects and performed a multi-dimensional comparative analysis. CT, MRI was performed in Resting state-processed and reported by neuro radiologists A 45 mins EEG procedures were used using all the activation procedures such as awake state, sleep deprived, sleep state, eye closure, eye opening, hyperventilation, photogenic stimulation.

## Results

Most common age group we found in our study was [10-20) years, In that group, 41persons/35.04% were affected. Following age group is [20 -30), in that group, 28 persons/ 23.93% were affected and in [0 -10) age group, 16 persons / 13.68% and in [30 -40) age group, 14 persons /11.97% and in [40-50) age group, 11 persons/9.40% in [50 -60) age group, 3 persons/2.56% and finally in [70 -80) age group, we found only one person/0.85% was affected. So the mean affected age is 23 years with standard deviate on (SD) of 14 years.

Males were commonly affected more by 66% in this study.(no need other info) Aura focality were analysed by history collected from the patients. We analysed how it started, nature of onset, environment during onset, duration of aura, previous history, its progress and interpretation [Figure 1].

**Figure 1:**
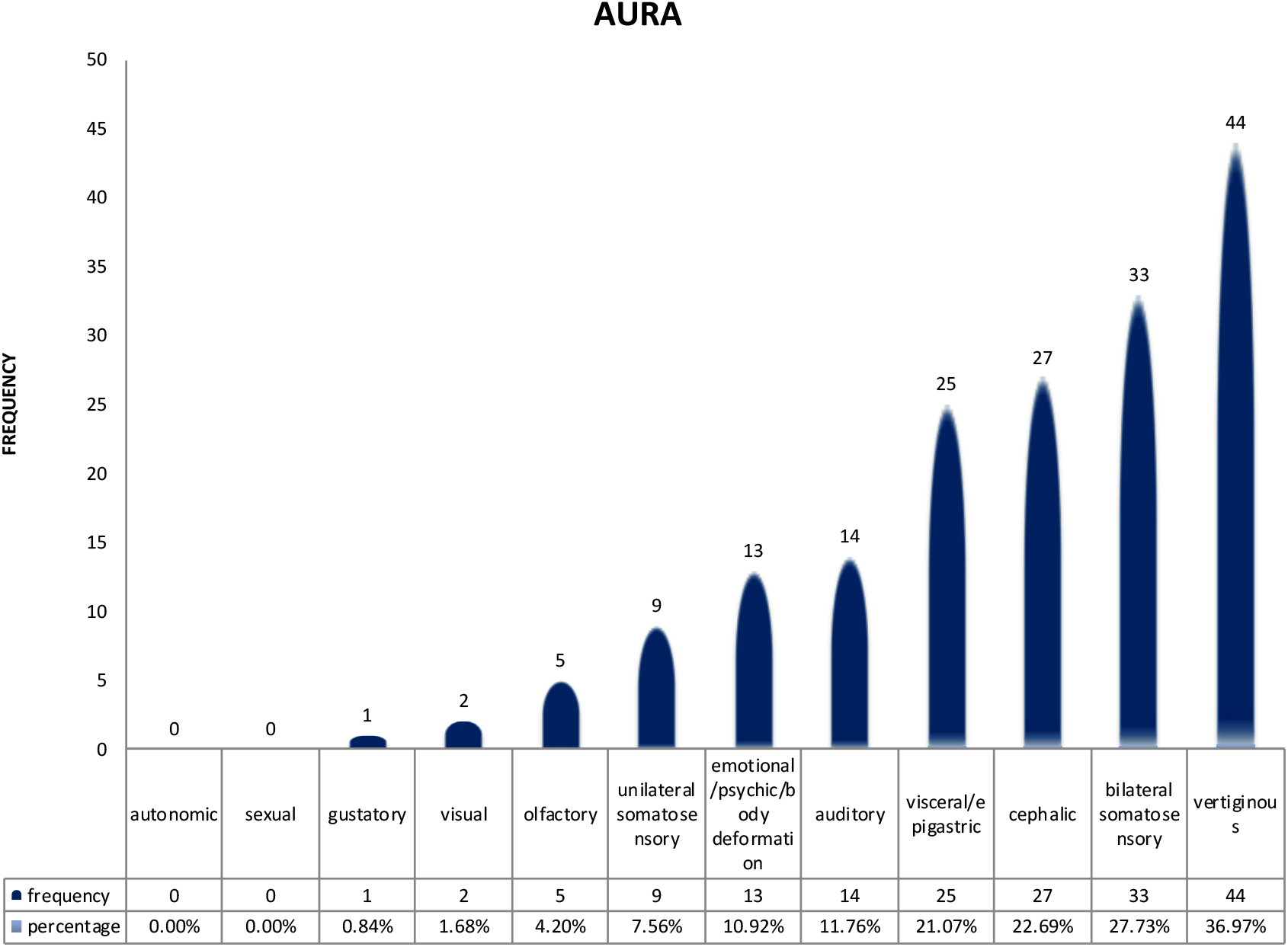
Histogram of the nature of aura in our subjects

While analyzing the presence of Aura in the patients we found the most common aura is vertiginous category having 44 patients / 36.97%. Next common aura found was somato sensory / bilateral in 33 patients / 27.73%. Third aura found in our study was cephalic in 27 patients / 22.69%. Fourth aura found was Visceral /epigastric in 25 persons / 21.07%. Fifth aura was Auditory in 14 patients / 11.76%. And the other following auras were found in decreasing frequencies with percentage such as-Olfactory aura 5 patients / 4.20%. Visual aura 2 patients / 1.68%. Gustatory aura – 1 person / 0.84%. In our study, No Sexual and Autonomic auras were reported in patients.

While analyzing the most common automatism in our study, Gestural/Manual type automatism was found in 72 patients / 60.50%. Vacant starring type in 67 patients / 56.30%. Oroalimentary type in 58 persons / 48.74%. Verbal type in 41 patients / 34.45%. Ambulatory type in 41 patients / 34.45%. Eye blinking type in 17 patients / 14.29%. Behavioral arrest in 16 patients / 13.45%. Running type in 14 patients / 11.76%. and other Automatism types of Fumbling, Sexual/Genital, Mimetic, Backward walking, Circling were found in around 5% of patients examined [Figure 2].

**Figure 2:**
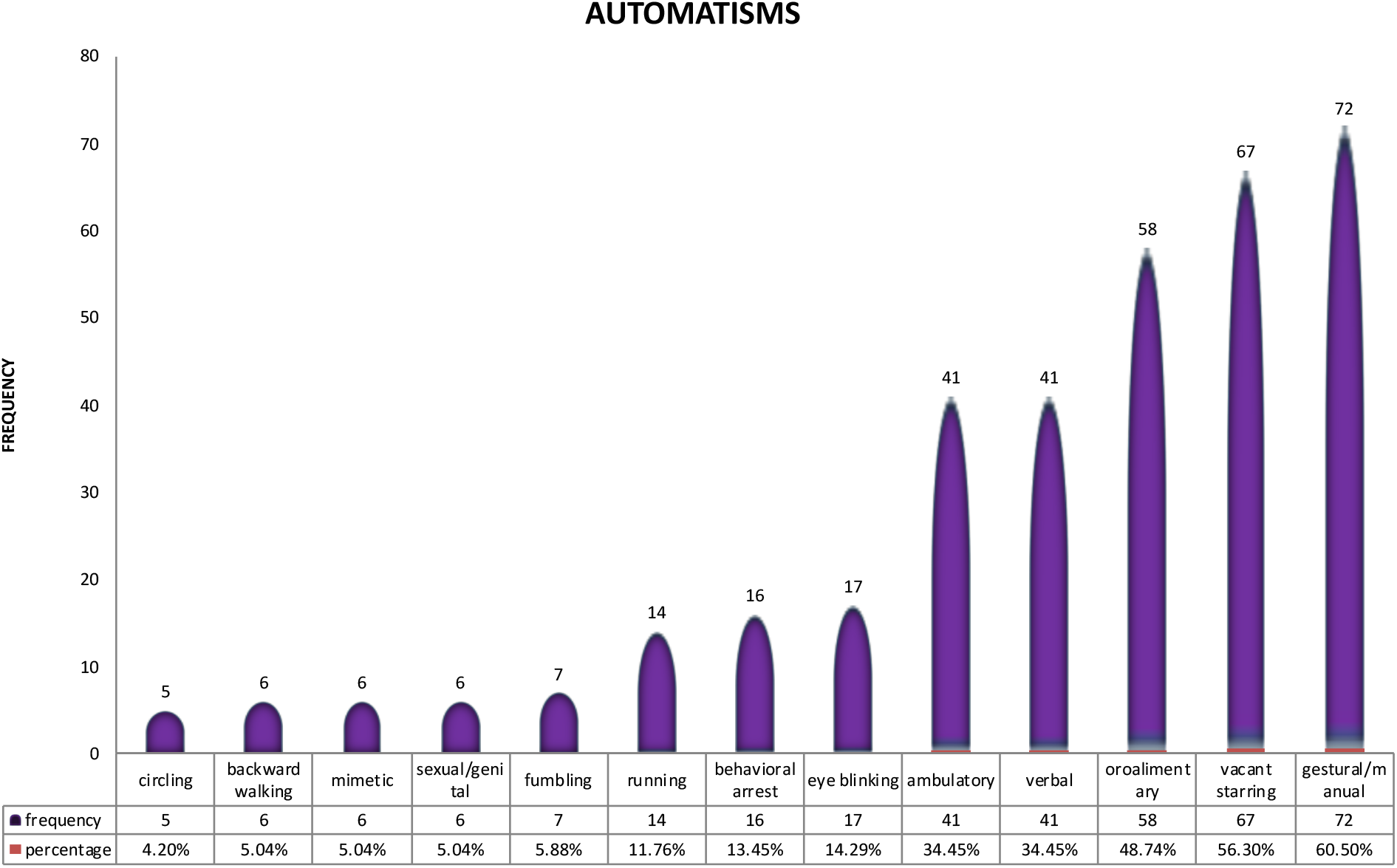
Histogram of the nature of automatisms in our subjects

A study on Seizures of Temporal lobe origin / temporal lobe epilepsy showed that [40-80 %) of patients had oral & manual automatism, whereas Young children with focal seizures of Temporal lobe had pre-dominantly behavioral arrest with unresponsiveness and children of age group 5 to 6 years showed Oroalimentary followed by Oral and Gestural automatism.

The studies on Seizures of Frontal lobe origin / frontal lobe epilepsy showed that motor automatisms such as involving the lower extremities (pedaling or bicycling movements) while Sexual automatisms and vocalizations were prominent.

### Past history

And in our study, 42 Patients / 34.43% were progressed to GTCS. In the Post ictal period, 18 patients / 20.17% had confusion, 5 patients / 0.84% had aggressiveness, 1 patient / 0.84% was with Amnesia for one day, 1 patient / 0.84% drinks water and 1 patient / 0.84% had Todds palsy.

Significant number of patient’s Past history had Febrile seizures (21 patients / 17.65%) History of Hie/mr in 8 patients / 6.72% and 5 persons / 4.20% each had brain fever, head injury, neoatal seizures and ADHD in 2 persons / 1.68%.

On analyzing the duration of aura, we found that most frequently the duration of aura was lasted for only 5 mins in 25 patients / 20.49% and maximum duration of aura was found as 30 mins in 11 patients / 9.02% [Figure 3].

**Figure 3:**
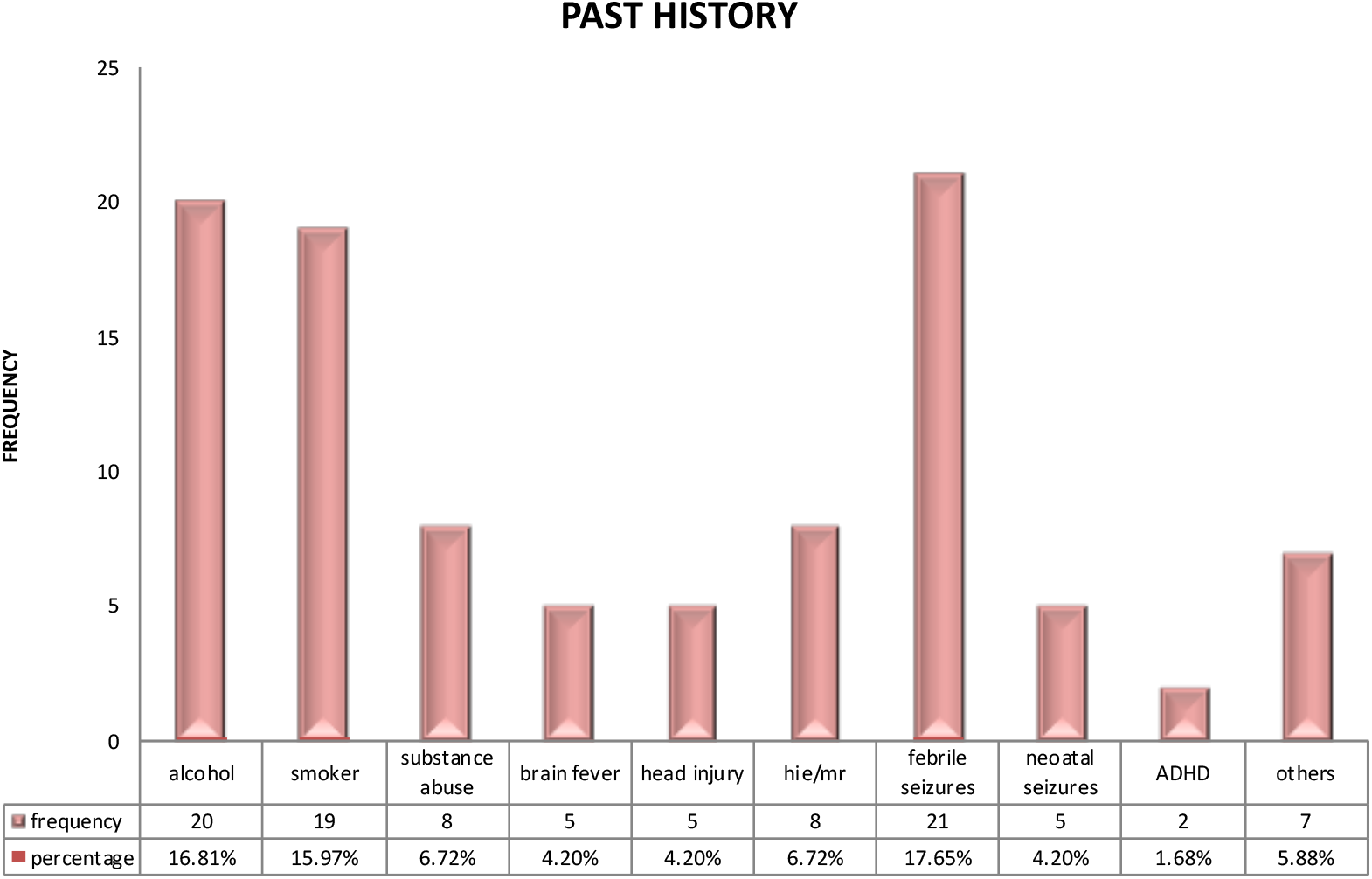
Histogram of past history in our subjects.

### Electroencephalography

EEG was found to be abnormal with epileptiform activity such as spike and wave discharges and some non epileptiform activities such as focal or diffuse slowing or focal spikes in 50 patients / 42.37%.

In this study, temporal lobe epilepsy was found to be more common in frequency, next to which was frontal lobe epilepsy. In temporal lobe epilepsy the most common EEG abnormality was found as non-epileptiform activity in 12 patients followed by Focal sharp /spikes-temporal origin in 6 patients and Focal spikes / sharp extra temporal origin in 3 patients and bilateral epileptiform activity in 2 patients. In frontal lobe epilepsy, the most common EEG abnormality was found as bilateral epileptiform activity in 8 patients followed by non-epileptiform activity in 5 patients and focal sharp/spikes temporal origin in 4 patients and NO person was found EEG abnormality in Focal spikes /sharp extra temporal origin [Figure 4].

**Figure 4:**
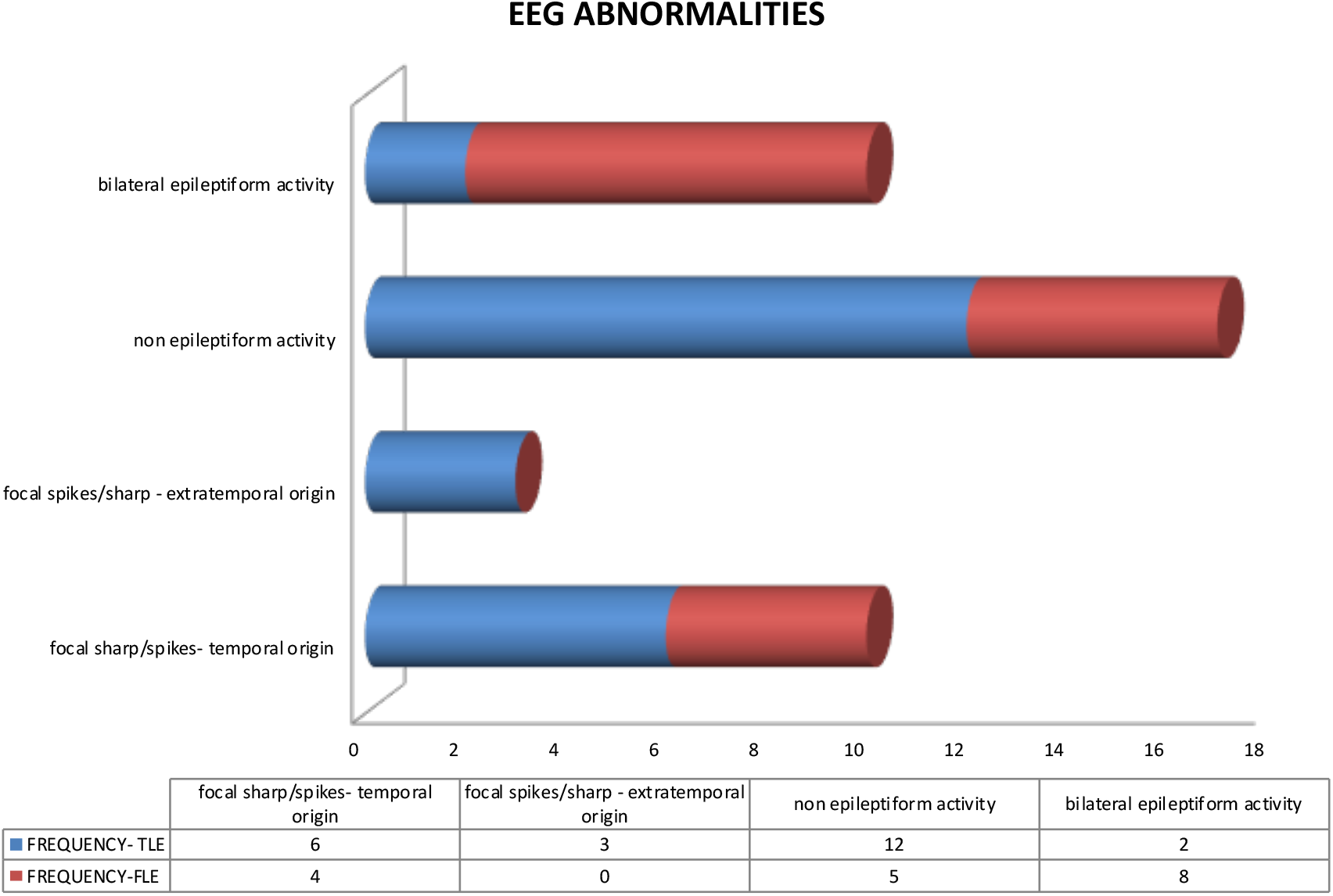
Histogram on EEG abnormalities in our subjects

Our study show temporal lobe epilepsy was the most common type of epilepsy in 69 patients / 58.47%, followed by frontal lobe epilepsy in 36 patients / 30.51%. As per our study, In temporal lobe epilepsy, 23 out of 69 patients which is 33.33% had abnormal EEG and in frontal lobe epilepsy, 17 out of 36 patients which is 47.22% had abnormal EEG.

### Magnetic Resonance Imaging

Most Common MRI findings were Normal followed by Medial Temporal Lobe Sclerosis followed by traumatic gliosis with the percentages as described below. In our study 45.76% of patients had symptomatic seizures, whose MRI findings had predominantly MTLS/Hippocampal volume loss in 22 patients / 18.64% followed by Traumatic Gliosis in 15 patients / 12.71% CVA with Gliosis in 11 patients / 9.32%, ICSOL in 9 persons / 7.63% HIE in 6 patients / 5.08% and malformation in 3 patients / 2.54% [Figure 5]. We found that Temporal Lobe Epilepsy had Symptomatic Lesions in 35 out of 69 patients which is 50.72% and Frontal Lobe Epilepsy had symptomatic Lesions in 17 out of 36 patients which is 47.22%

**Figure 5:**
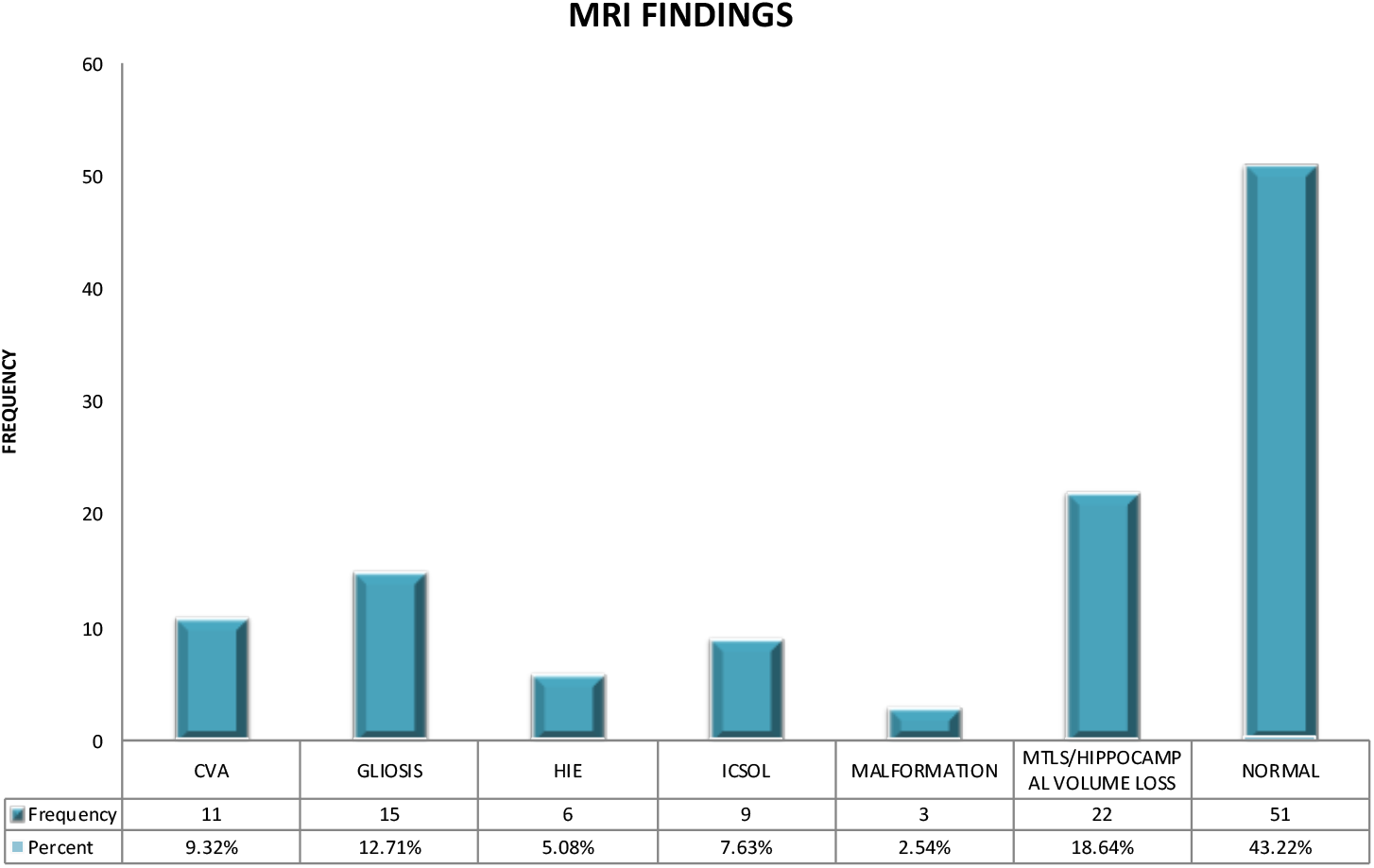
Histogram on MRI findings in our subjects.

We also found some lateralizing features: Unilateral ictal clonic activity or ictal dystonia suggests lateralization of the seizure to the contralateral hemisphere. Early forced head version suggests lateralization to the hemisphere contralateral to the direction of the head version i.e. if the head turns to the right, the seizure onset is in the left hemisphere. Ictal speech lateralizes to the non-dominant hemisphere. Ictal aphasia lateralizes to the dominant hemisphere.Post-ictal nose-wiping lateralizes to the hemisphere ipsilateral to the hand used for nose-wiping. Unilateral eye-blinking lateralizes to the hemisphere ipsilateral to the eye-blinking. Ictal vomiting lateralizes to the non-dominant hemisphere.

## Discussion

One of the most controversial issues in epilepsy research is whether prolonged febrile seizures cause mesial temporal sclerosis and temporal lobe epilepsy. Retrospective studies from tertiary epilepsy centers report that many adults with intractable epilepsy have a history of prolonged or atypical febrile seizures in childhood. However, population-based studies have failed to confirm this association, as have prospective studies of febrile seizures. Our study focused on complex partial seizure patients in Thanjavur, India, confirms the association between atypical febrile seizures and complex partial seizures.

Mesial temporal sclerosis probably has different causes. A number of retrospective studies showed that complex febrile seizures are a causative factor for the later development of mesial temporal sclerosis and temporal lobe epilepsy. The association between febrile seizures and temporal lobe epilepsy probably results from complex interactions between several genetic and environmental factors.

Some earlier studies that used EEG and MRI can be used as a biomarker for prediction have projected around 12% develops epilepsy 8-12 years later-according to studies^9^ in contrast to 18% in our study.

While analyzing the presence of Aura in the patients we found the most common aura is vertiginous category whereas another similar study^8^ on 2013 projected patients show 19 out of 57% falls in this category and a study^10^ on 1359 patients show results as 12 out of 56% fall in this category. Next common aura found was somato sensory in comparison to other studies such as Gowers et al^8^ suggests 18 out of 57% and Lennox et al^10^ suggests 8.5% out of 56%. Third aura found in our study was cephalic in contrast to 8 out of 57%^8^ and in 5 out of 56%^10^. Fourth aura found was Visceral /epigastric compared to 18 out of 57%^8^ and 14.5 out of 56%^10^. And the fifth aura was Auditory in contrast to 6 out of 57%^8^ and 2 out of 56%^10^. Sixth aura found was emotional /psychi /body deformation whereas in 8 out of 57%^8^ and study 11 out of 56%^10^. And the other following auras were found in decreasing frequencies with percentage such as-Olfactory aura (1%^8^-1%^10^) Visual (16%^8^-6.5%^10^) Gustatory aura (1.5%^8^ and 0.1%^10^) In our study, No Sexual and Autonomic auras were reported in patients.

Some earlier studies^11–13^ suggests lateralising Auras of focal epilepsies such as Somatosensory aura lateralised are more common in Parietal lobe (61%) followed by Frontal lobe (17.5%). Epigastric/emotional Aura lateralised in Temporal lobe (52%) followed by 12.5% in Frontal lobe. Cephalic aura lateralized in Frontal lobe (12.5%). Then, Psychic aura lateralized in two lobes equally, Temporal lobe(15%) and Parietal lobe (15%). Visual aura lateralized mostly in Occipital lobe (74%) followed by Temporal (11%). Auditory, Olfactory, Gustatory – all 3 Auras were lateralized mainly in Temporal lobe only (11% each). Vertiginous aura lateralized in Temporal lobe (11%) followed by Parietal lobe (9%).

Automatisms and aura correlated and is consistent with earlier investigations: 41.5 % in temporal lobe epilepsy^12^, 32 % in Extra temporal lobe epilepsy. Various studies showed that temporal intermittent rhythmic delta activity is found in up to 28% of patients evaluated for temporal lobe epilepsy, bi-temporal sharp-wave foci are noted in 25% to 33% of patients and on a single routine EEG recording, 30% to 40% of patients had normal Interictal findings; activating techniques can reduce this to approximately 10%. In 10% to 30% of patients with frontal lobe epilepsy, they observed EEG activity of bifrontal spike – and – wave.

Altogether our results on aura progression and nature, their lateralization, nature of automatisms, were consistent with the earlier studies as above^8,10–13^.

We believe this study summarizes the complex partial seizure features, origin, and their link to MTS in our subject pool from Thanjavur, India. There are some limitations to our study, especially with no video EEG monitoring and no invasive EEG recording. We aim to improve them in our future studies.

## Data Availability

On request to the author

